# Strength and durability of antibody responses to BNT162b2 and CoronaVac

**DOI:** 10.1101/2022.02.11.22270848

**Authors:** Benjamin J. Cowling, Irene O. L. Wong, Eunice Y. C. Shiu, Amber Y. T. Lai, Samuel M. S. Cheng, Sara Chaothai, Kelvin K. H. Kwan, Mario Martín-Sánchez, Leo L. M. Poon, Dennis K. M. Ip, Gabriel M. Leung, Nancy H. L. Leung, J. S. Malik Peiris

## Abstract

We studied 2780 adults in Hong Kong who received CoronaVac inactivated virus vaccine (Sinovac) and BNT162b2 mRNA vaccine (“Comirnaty”, BioNTech/Fosun Pharma). We found stronger and more durable antibody responses to two doses of the mRNA vaccine, and slightly stronger initial antibody responses to each vaccine in younger adults and women.

## INTRODUCTION

Vaccines for coronavirus disease 2019 (COVID-19) provide protection against infection and can also ameliorate disease severity if breakthrough infections occur [1]. Protection against infection is likely mediated through antibodies, and the effectiveness of different vaccine technologies against mild symptomatic infection has been shown to correlate with the strength of neutralizing antibody responses to those vaccines [2-4]. In particular, antibody responses after receipt of inactivated COVID-19 vaccines have been shown to be somewhat weaker than antibody responses to mRNA vaccines [5, 6], although T cell responses are more comparable between the two vaccine technologies [6]. There are relatively fewer studies comparing rates of antibody waning over time, particularly with inactivated vaccines [7, 8].

In Hong Kong, two vaccines have been made available to the general public through a mass vaccination program since early 2021, the CoronaVac inactivated virus vaccine (Sinovac) and the BNT162b2 mRNA vaccine (“Comirnaty”, BioNTech/Fosun Pharma). As of 11 February 2022, 11.7 million vaccine doses have been administered, with 74% of the population of about 7.4 million residents receiving at least one vaccine dose, 67% receiving at least two doses, and 16% receiving three doses. Of the 11.7 million doses administered to date, 4.5 million were CoronaVac and 7.2 million were BNT162b2. Here, we followed up vaccinated individuals in Hong Kong to study the strength and durability of antibody responses to vaccination for up to six months after the second dose.

## METHODS

### Recruitment and Follow-up of Participants

We analyzed data from two ongoing community-based cohort studies of infections and immunity. In the “<u>E</u>valuating <u>P</u>opulation Immunity in <u>H</u>ong <u>K</u>ong” (EPI-HK) cohort study which was initially designed to study influenza and later also expanded to COVID-19, we enrolled individuals from the community starting in July 2020, and included individuals regardless of COVID-19 vaccination status. For analyses reported here we include the subset of study participants who were aged ≥18y and have received two doses of COVID-19 vaccination. Blood samples are collected from each participant at approximately six-month intervals, with additional blood samples collected one month after any dose of COVID-19 vaccination.

In the “<u>CO</u>VID-19 <u>VA</u>ccine <u>R</u>esearch” (COVAR) cohort study which aims to study vaccine immunogenicity and long-term antibody responses in COVID-19 vaccine recipients, we enrolled adults ≥18y of age from the general community and from Community Vaccination Centres. In the initial protocol we aimed to enroll individuals shortly before receiving the first dose of any COVID-19 vaccine in Hong Kong, this was later relaxed to allow enrollment of individuals after receiving one or more vaccine doses. Blood samples are collected from each participant at baseline, one month after any dose of COVID-19 vaccination, and at 6, 12, 24 and 36 months after the second dose of vaccination.

Similar data are collected from participants in both studies, including information on demographics and socio-economic status, underlying medical conditions and regular medication use, and the type and dates of any COVID-19 vaccine doses received. Participants receive coupons worth HK$100 (approximately US$13) at each blood draw.

### Ethics

Written informed consent was obtained from all participants. The study protocols were approved by the Institutional Review Board of the University of Hong Kong.

### Laboratory Methods

Sera were extracted from the clotted blood samples within 24 hours after collection, divided into 2-4 aliquots, and stored at -80°C until subsequent serologic testing. Samples were tested with an ELISA for spike receptor binding domain and a surrogate virus neutralization test (sVNT) as previously described [9]. We previously demonstrated that both assays can be used as proxy for neutralizing antibody against SARS-CoV-2 as measured by plaque reduction neutralization test with live virus [10].

We obtained sVNT kits from GeneScript USA, Inc, New Jersey and the tests carried out according to the manufacturer’s instructions. The % inhibition of each serum was calculated as Inhibition (%) = (1 - Sample OD value/Negative Control OD value) x100. Inhibition (%) of ≥30% was regarded as a positive result while <30% was considered negative [9].

ELISA was carried out as previously described [9]. 96-well ELISA plates (Nunc MaxiSorp, Thermo Fisher Scientific) were coated overnight with 100 ng per well of the purified recombinant RBD protein in PBS buffer. The plates were then blocked with 100 μl of Chonblock blocking/sample dilution ELISA buffer (Chondrex Inc, Redmon, US) and incubated at room temperature for 2 h. Each serum or plasma sample diluted 1:100 in Chonblock blocking/sample dilution ELISA buffer was added to duplicate wells and incubated for 2 h at 37°C. After washing with PBS containing 0.1% Tween 20, horseradish peroxidase (HRP)-conjugated goat anti-human IgG (1:5,000, GE Healthcare) was added for 1 h at 37°C. The ELISA plates were then washed in PBS with 0.1% Tween 20. Subsequently, 100 μL of HRP substrate (Ncm TMB One; New Cell and Molecular Biotech Co. Ltd, Suzhou, China) was added into each well. After 15 min incubation, the reaction was stopped by adding 50 μL of 2 M H_2_SO_4_ solution and analyzed on a Sunrise (Tecan, Männedorf, Switzerland) absorbance microplate reader at 450 nm wavelength. Optical density above 0.5 was considered a positive result.

### Statistical Analysis

We assessed mean antibody levels measured by ELISA (optical density, OD) and sVNT (% inhibition) after the first and second dose of COVID-19 vaccination. It was recommended that individuals should receive their second dose 21 and 28 days after the first dose for BNT162b2 and CoronaVac, respectively. We did not include any samples collected after receipt of a third dose, or any samples collected after a confirmed SARS-CoV-2 infection. Differences in antibody levels between vaccine type overall or within each category of age or sex were compared using t-tests. To evaluate the duration after which antibody level fall below the manufacturer’s defined sVNT threshold of seropositivity at 30% inhibition (Supplementary Information), we fitted a smoothing spline to antibody levels over time starting at 14 days and up to 180 days after receipt of the second dose. Statistical analyses were conducted using R version 4.1.2 (R Foundation for Statistical Computing, Vienna, Austria).

## RESULTS

We included data from 2780 vaccinated individuals including 2172 from the EPI-HK study and 608 from the COVAR study who had provided at least one post-vaccination blood sample by 23 December 2021, contributing 1528 blood samples collected from 799 individuals who received CoronaVac (median age 59.4 years, 46.6% male), and 3939 blood samples collected from 1981 individuals who received BNT162b2 (median age 49.3 years, 44.7% male). Additional information on the demographics of study participants is provided in Appendix Table 1.

Antibody levels increased to moderate levels following two doses of CoronaVac (Figure 1A) compared to very high levels following two doses of BNT162b2 (Figure 1B). The mean sVNT antibody levels measured against ancestral virus at 14-42 days after the second dose for CoronaVac were 53.7% (95% confidence interval, CI: 51.2, 56.3) and for BNT162b2 were 94.3% (95% CI: 93.8, 94.7), with significant differences between the two vaccines overall and within each age group (Appendix Table 2). For both vaccines, the observed post-vaccination levels had a slight inverse correlation with age, and average levels were slightly higher in females than in males (p-values <0.01 for these within-group comparisons) (Appendix Table 2). Overall, antibody levels remained below the threshold of 30% after the second dose in 23% and 0.5% of participants who received CoronaVac and BNT162b2, respectively. In particular, the antibody levels did not reach the threshold of 30% in 51% of older adults ≥60 years old who received CoronaVac vaccination.

**Figure 1:**
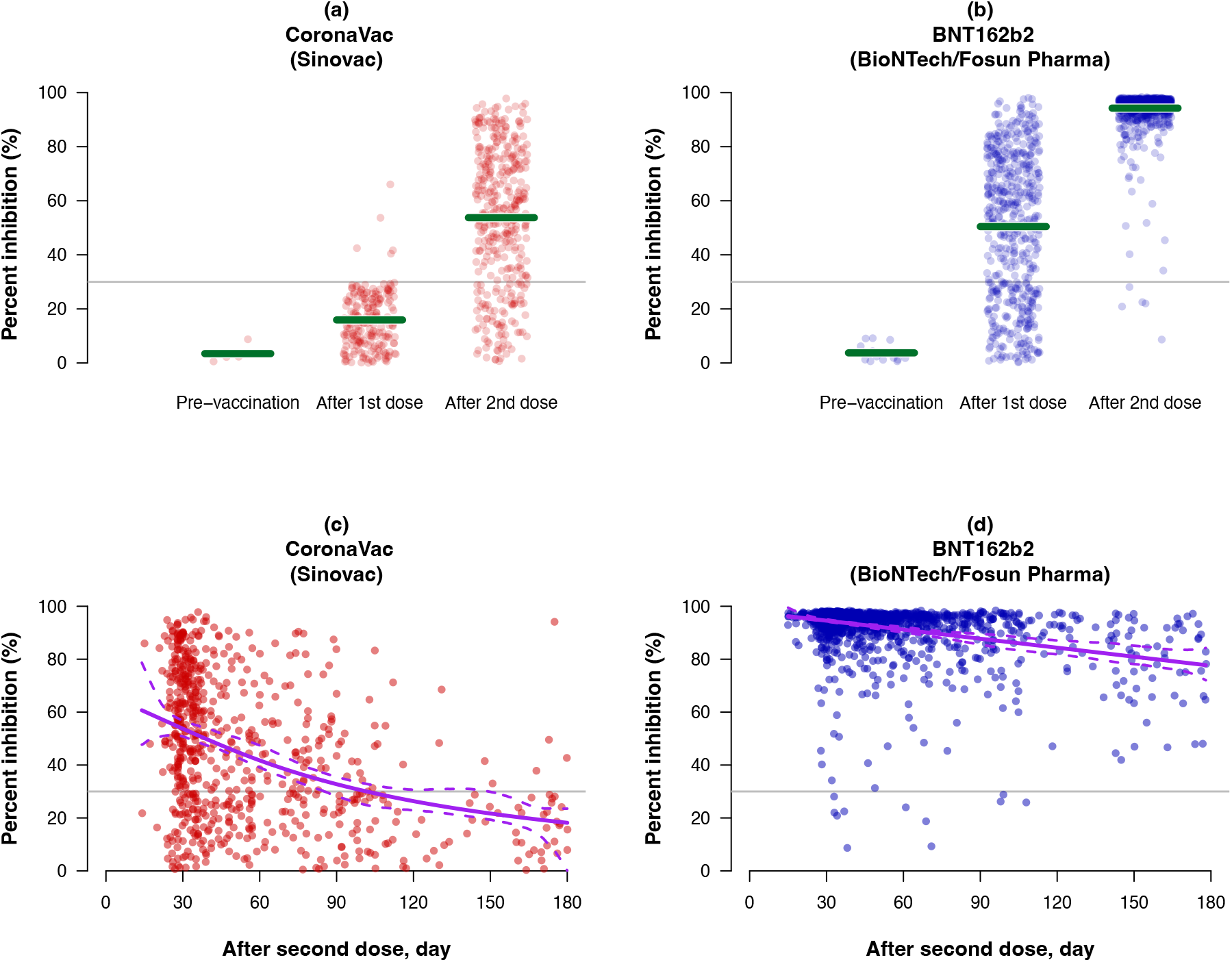
Post-vaccination rises followed by longer-term decay in neutralizing antibody levels against the ancestral SARS-CoV-2 virus after receipt of two doses of CoronaVac or BNT162b2 vaccination, measured by a surrogate virus neutralization test. Panels A and B show antibody levels prior to receipt of vaccination (“Pre-vaccination”), 14-35 days after receipt of the first dose but no later than the day of receipt of second dose (“After 1^st^ dose”), and 14-42 days after receipt of the second dose (“After 2^nd^ dose”), for an inactivated (CoronaVac) or mRNA (BNT162b2) vaccine respectively. Green lines represent mean % inhibition in the respective time periods. Panels C and D show antibody levels between 14- 180 days after the receipt of a second dose of CoronaVac or BNT162b2, respectively. Solid purple lines represent the spline fit and dotted lines the associated 95% pointwise bootstrap confidence intervals for the estimated changes over time in mean antibody levels. In all four panels, the grey horizontal line indicates the manufacturer’s defined threshold for seropositivity at 30% inhibition.

Antibody levels started to decline gradually over time starting from one month after recipient of the second dose for both vaccines, with faster antibody waning in the recipients of CoronaVac (Figure 1C and 1D). In recipients of CoronaVac, the fitted curve for average antibody level fell below the threshold of 30% by around 4 months after second dose, while remaining elevated for at least six months in recipients of BNT162b2. Consistently, we continued to observe significant differences in antibody levels 4-6 months after vaccination between the two vaccines overall and within each age group (Appendix Table 3). The mean sVNT antibody levels at 120-180 days after the second dose overall were 21.2% (95% CI: 16.9, 25.6) and 81.1% (95% CI: 78.0, 84.3) for CoronaVac and BNT162b2 respectively.

Specifically, sVNT antibody levels fell to below the threshold of 30% in 84% of participants who received CoronaVac, versus none among those who received BNT162b2. However, with a smaller sample size here we were not able to identify statistically significant differences by age or sex within each vaccine group. We also examined antibody levels by ELISA, finding similar comparative patterns of boosting and waning between the two vaccine types (Appendix Figure 1).

## DISCUSSION

Evidence on antibody boosting and waning after vaccination is important as it affects the potential timing of subsequent booster vaccine doses. Here, we demonstrate higher levels of antibody to SARS-CoV-2 following two doses of the mRNA vaccine BNT162b2 compared to two doses of the inactivated vaccine CoronaVac [5, 6], and longer durability of responses for BNT162b2 for at least six months following the receipt of a second dose [7, 11]. Our finding of rapid waning of antibody following receipt of CoronaVac echoes previous reports [7, 11] and indicates that a third dose would likely be needed sooner, and indeed in Hong Kong third doses have been recommended at ≥3 months after the second dose of CoronaVac. It is important to recognize, however, that protection against severe COVID could be retained even in spite of waning of protection from infection [12].

We found stronger responses to two doses of both vaccines in females and in younger adults, consistent with other studies. For example Uwamino et al reported similar observations in a cohort of 673 recipients of BNT162b2 [13], while Xu et al. reported stronger and more durable antibody responses in females and younger recipients of an inactivated vaccine [14]. There are a number of potential underlying biological mechanisms that could lead to a sex difference, and stronger antibody responses in women have been reported for other vaccines [15].

There are a number of limitations of our study. First, we did not measure neutralizing antibody against live SARS-CoV-2 but used a surrogate neutralization assay which has a high correlation with live virus neutralization titers [16]. We did not assess antibodies against variants such as Delta or Omicron, but expect antibody levels to be reduced against variants as reported in other studies [16, 17]. We did not assess T cell responses which could contribute to protection against severe disease and could be less affected by variants [18].

In conclusion, we identified weaker boosting and faster waning of antibodies against SARS- CoV-2 in recipients of CoronaVac compared to BNT162b2, and would expect this to correlate with lower levels of protection against symptomatic infection for CoronaVac. In individuals of any age or sex, BNT162b2 responses were stronger and more durable than responses to CoronaVac. The weaker and less durable responses following CoronaVac support earlier provision of third doses to persons who previously received two doses of this vaccine.

## Data Availability

All data produced in the present study are available upon reasonable request to the authors

## ACKNOWLEDGMENTS

We acknowledge colleagues including Jemmi Ho, Joyce Lau, Shadow Lau, Leslie Leung (EPI-HK study), Alex Chan, Vincent Fung, Raina Kwan, Lilly Wang, Benny Wong (COVAR study) and other part-time colleagues for technical support in preparing and conducting this study and enrolling participants; Anson Ho for setting up the database; Leo Luk, Karl Chan, John Li, Yonna Leung, Leo Tsang and Zacary Chai for laboratory support; Hetti Cheung, Victoria Wong, Bobo Yeung at HKU Health System; and King-man Ho, Cindy Man and other colleagues at the Community Vaccination Centres at Ap Lei Chau Sports Centre, Gleneagles Hospital and Hong Kong Central Library for facilitating the two cohort studies.

## FINANCIAL SUPPORT

This project was financially supported by the Health and Medical Research Fund of the Food and Health Bureau of the Hong Kong SAR Government (project nos. COVID1903001 and COVID190126), the Theme-based Research Scheme (project no. T11-712/19-N) from the Research Grants Council from the University Grants Committee of Hong Kong, and the Wellcome Trust (grant number: 221013/Z/20/Z). BJC is supported by an RGC Senior Research Fellowship (grant number: HKU SRFS2021-7S03).

## POTENTIAL CONFLICTS OF INTEREST

BJC consults for AstraZeneca, Fosun Pharma, GSK, Moderna, Pfizer, Roche and Sanofi Pasteur. The authors report no other potential conflicts of interest.

---

**Appendix Figure 1:**
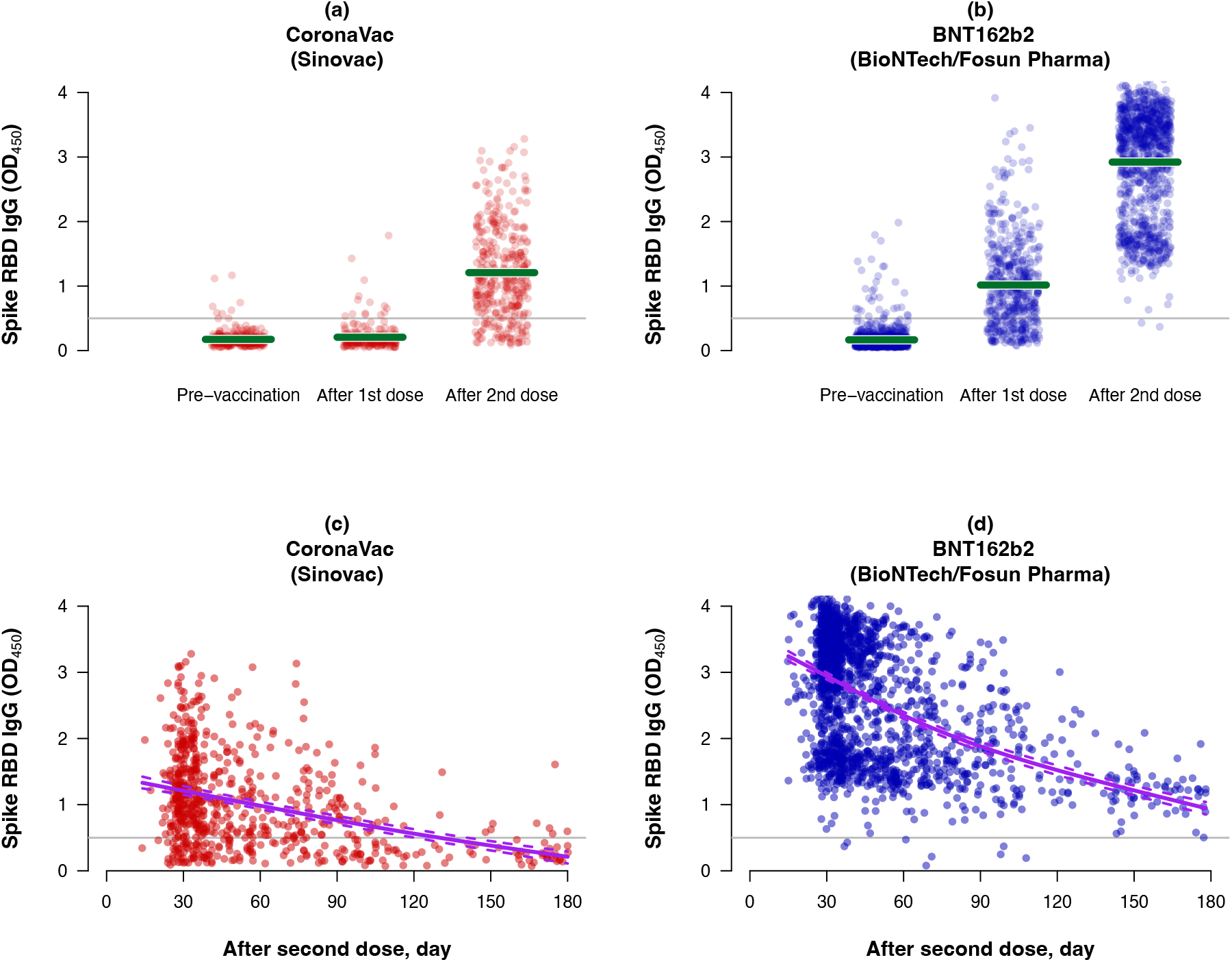
Post-vaccination rises followed by longer-term decay in IgG antibody levels against the ancestral SARS-CoV-2 virus after receipt of two doses of CoronaVac or BNT162b2 vaccination, measured by an ELISA assay against the receptor binding domain of the spike protein. Panels A and B show antibody levels prior to receipt of vaccination (“Pre-vaccination”), 14-35 days after receipt of the first dose but no later than the day of receipt of second dose (“After 1^st^ dose”), and 14-42 days after receipt of the second dose (“After 2^nd^ dose”), for an inactivated (CoronaVac) or mRNA (BNT162b2) vaccine respectively. Green lines represent mean optical density (OD) in the respective time periods. Panels C and D show antibody levels between 14-180 days after the receipt of a second dose of CoronaVac or BNT162b2, respectively. Solid purple lines represent the spline fit and dotted lines the associated 95% pointwise bootstrap confidence intervals for the estimated changes over time in mean antibody levels. In all four panels, the grey horizontal line indicates the manufacturer’s defined threshold for seropositivity at OD of 0.5.

**Appendix Table 1.**
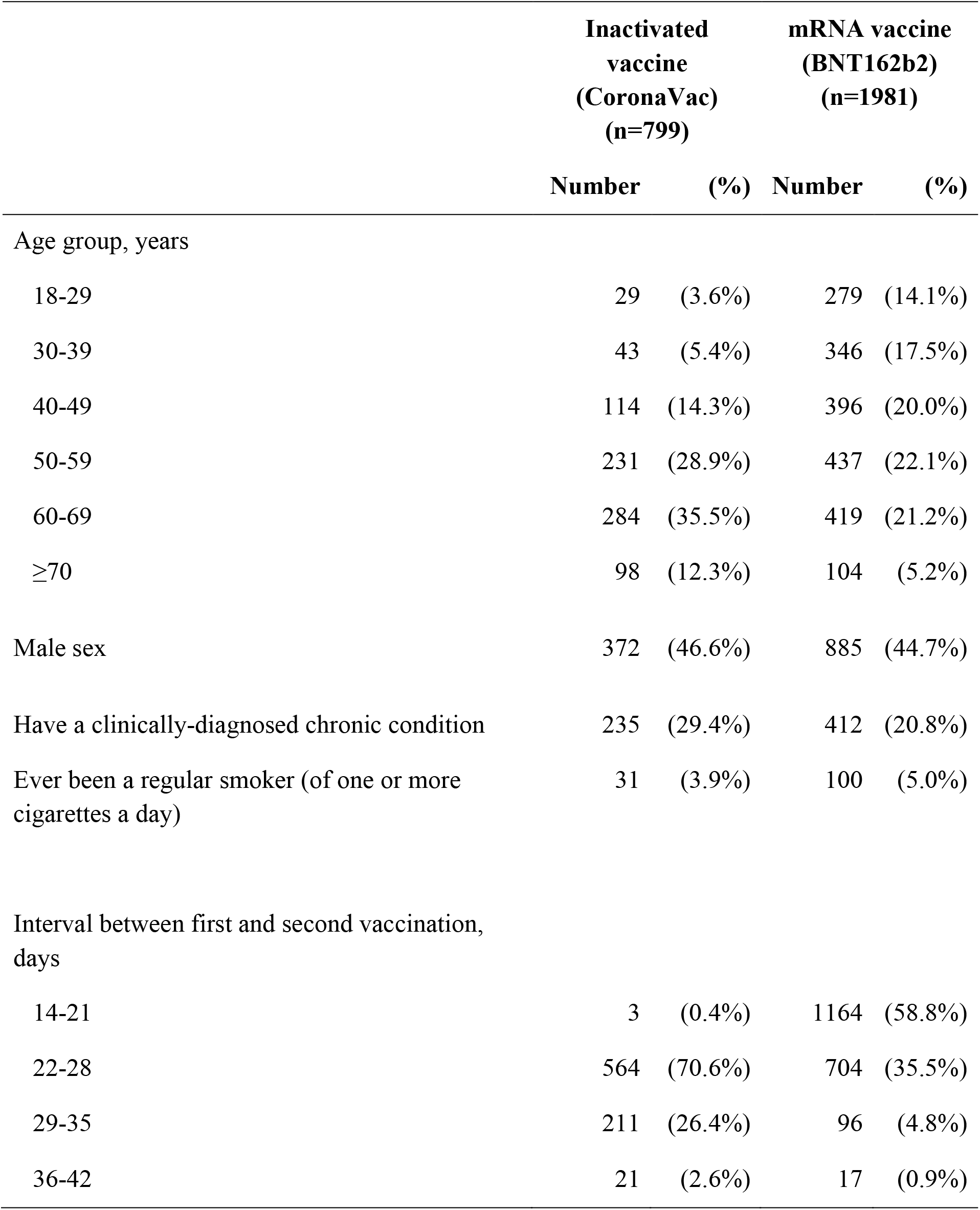
Characteristics of study participants who received two doses of inactivated vaccine (CoronaVac) or mRNA vaccine (BNT162b2) (n=2780). Participants from both the EPI-HK and COVAR studies who have antibody levels measured at least once between 14-180 days after recipient of the second dose are included.

**Appendix Table 2.** Mean sVNT levels measured against ancestral SARS-CoV-2 virus at 14-42 days after the receipt of the second vaccine dose of an inactivated (CoronaVac) or mRNA (BNT162b2) vaccine, overall and by age and sex (n=1431). Only participants from both the EPI-HK and COVAR studies who have antibody levels measured between 14-42 days after recipient of the second dose are included.

| Group | Inactivated vaccine<br>(CoronaVac)<br>(n=378) |  | mRNA vaccine<br>(BNT162b2)<br>(n=1053) |  | p-value* |
| --- | --- | --- | --- | --- | --- |
|  | sVNT | (95% CI) | sVNT | (95% CI) |  |
| Overall | 53.7 | (51.2, 56.3) | 94.3 | (93.8, 94.7) | <0.001 |
| Age group,<br>years |  |  |  |  |  |
| 18-29 | 69.6 | (58.8, 80.4) | 96.5 | (96.2, 96.8) | <0.001 |
| 30-39 | 54.0 | (44.1, 63.9) | 94.7 | (93.7, 95.6) | <0.001 |
| 40-49 | 53.1 | (46.1, 60.2) | 94.4 | (93.6, 95.3) | <0.001 |
| 50-59 | 56.9 | (52.5, 61.3) | 94.0 | (93.2, 94.9) | <0.001 |
| 60-69 | 51.6 | (46.8, 56.4) | 93.4 | (92.1, 94.6) | <0.001 |
| ≥70 | 46.3 | (38.9, 53.6) | 92.3 | (89.6, 94.9) | <0.001 |
| Male | 48.0 | (43.9, 52.0) | 93.3 | (92.5, 94.2) | <0.001 |
| Female | 58.6 | (55.4, 61.8) | 95.0 | (94.5, 95.4) | <0.001 |

\*p-values comparing antibody levels between the two vaccines overall and in each subset of participants (i.e. each row of this table) were estimated by t-tests.

**Appendix Table 3.** Mean sVNT levels measured against ancestral SARS-CoV-2 virus at 120-180 days after the receipt of the second vaccine dose of an inactivated (CoronaVac) or mRNA (BNT162b2) vaccine, overall and by age and sex (n=134). Only participants from both the EPI-HK and COVAR studies who have antibody levels measured between 120-180 days after recipient of the second dose are included.

| Group | Inactivated vaccine<br>(CoronaVac)<br>(n=51) |  | mRNA vaccine<br>(BNT162b2)<br>(n=83) |  | p-value* |
| --- | --- | --- | --- | --- | --- |
|  | sVNT | (95% CI) | sVNT | (95% CI) |  |
| Overall | 21.2 | (16.9, 25.6) | 81.1 | (78.0, 84.3) | <0.001 |
| Age group,<br>years<sup>†</sup> |  |  |  |  |  |
| 18-39 | -<sup>††</sup> | - | 85.8 | (80.8, 90.7) | <0.001 |
| 40-59 | 23.9 | (16.1, 31.6) | 79.3 | (74.3, 84.2) | <0.001 |
| ≥60 | 18.7 | (14.7, 22.8) | 78.9 | (72.1, 85.7) | <0.001 |
| Male | 16.5 | (13.4, 19.7) | 80.7 | (75.9, 85.6) | <0.001 |
| Female | 24.8 | (17.6, 32.0) | 81.5 | (77.3, 85.7) | <0.001 |

\*p-values comparing antibody levels overall and in each subset between the two vaccines (i.e. each row of this table) were estimated by t-tests.
<sup>†</sup>Due to smaller sample sizes we used broader age groups in this analysis compared to the analysis shown in Appendix Table 2.
<sup>††</sup>Not estimated

## Notes

### Author Declarations

The study protocols were approved by the Institutional Review Board of the University of Hong Kong

